# Spurious Correlation? A review of the relationship between Vitamin D and Covid-19 infection and mortality

**DOI:** 10.1101/2020.05.25.20110338

**Authors:** Arpita Srivastava, Vivek Kumar

## Abstract

The study reviews the evidence presented in a recent study linking vitamin D levels and Covid-19 infection and mortality. It was argued that correlation alone may not be useful in establishing a relationship between vitamin D levels and Covid-19 infections and mortality. Appropriate controls need to be included for improved understanding of the relationship. We proposed life expectancy as a potential control. Including this control in a regression model, we find that vitamin D levels are not a statistically significant predictor of Covid-19 infections or mortality. Life expectancy, on the other hand, was found to be statistically significant predictor of infections and mortality at country level.

## Introduction

Research community across the globe is intensely searching for a better understanding of Covid-19 in the hope of finding a cure or a vaccine. Any Covid-19 related potential treatment has met with a disproportionate response from the governments and society. For example, news of HCQ as a potential treatment led medical doctors writing prescriptions for themselves, deaths occurring, and diplomatic rows between countries. All this despite the fact that the scientific evidence behind HCQ was weak^1–3^.

A recent study found that vitamin D levels and Covid-19 infections and mortality are negatively correlated using a country level sample of 20 European countries^4^. Expectedly, the study was immediately shared across the world by news and social media. Altmetric data of the article shows that the study has been picked up by around 85 news outlets and has been tweeted thousands of times. News outlets stretched the conclusions of the study and claimed that vitamin D may reduce mortality risk by 50%. Such claims can lead people to overdosing on the vitamin which can have fatal consequences as has been the case with hydroxychloroquine (HCQ).

It is important to note that the study only found a statistically significant correlation between Vitamin D levels and Covid-19 cases and deaths. Drawing real life interpretation from correlations alone in secondary data is fraught with risk. A correlation between two variables must be controlled for other confounding variables to derive meaningful information^5^. In this study we re-test the relationship between vitamin D and Covid-19 infections and mortality by controlling for life expectancy. We find that the relationship not statistically significant. Life expectancy is a statistically significant predictor of both infections and mortality arising from Covid-19 at country level.

## Background of the study

Controlled experiments, such as clinical trials, are the gold standard of science. This is because competing explanations are controlled for. However, experiments are not always possible. This is almost always true in social sciences. In such contexts, research must depend on data already collected. Drawing inferences from secondary data is always fraught with risk of misinterpretation. For example, in a controlled experiment, a correlation between treatment and effect is most likely correct interpretation as competing explanations have been controlled for. However, a correlation in secondary data does not mean much unless the relationship is confirmed by further analysis.

Ilie and colleagues^4^ find that the Pearson correlation coefficient of mean vitamin D levels and Covid-19 cases and of mean vitamin D levels and Covid-19 deaths per million population is negative and statistically significant (r = − 0.4435, p < 0.05 and r = − 0.4378, p <0.05) based on data from 20 European countries. Does that mean vitamin D decreases risk of infections and mortality due to Covid-19? The correlation alone cannot be used to answer this question despite statistical significance. The correlation has the potential of being misleading as other factors have not been controlled for. Any factor that may impact both vitamin D levels and Covid-19 infections must be controlled for before arriving at any conclusion^5^.

One such factor might be life expectancy of the population of a country. A nation with higher life expectancy might have a more aging population than a nation with lower life expectancy. Age is a known risk factor of Covid-19^6^. Life expectancy and vitamin D levels too might be correlated. Older population is less exposed to sun – a source of vitamin D. Thus, we argue that differences in life expectancy may be correlated with Covid-19 infections and mortality on the one hand, and vitamin D levels on the other hand. By controlling for life expectancy, the true relationship between vitamin D and Covid-19 infections and mortality can be known.

## Method and Data

To test the true relationship between vitamin D levels and Covid-19 infections and mortality, we estimate the following two models which control for life expectancy:

CovidCases_i_ = β_1_*VitaminD_i_ + β_2_*LifeExpectancy_i_ + error_i_

CovidMortality_i_ = β_1_*VitaminD_i_ + β_2_*LifeExpectancy_i_ + error_i_

Where i represents countries in the sample. CovidCases are number of Covid-19 cases per million population reported in each country. CovidMortality is number of deaths attributed to Covid-19 per million population in each country. Vitamin D is Vitamin D(25)HD mean measured in nmol/L.

Data on Covid-19 Cases and mortality, and mean vitamin D levels is taken from Ilie and colleagues’ article^4^ so that the results are not influenced by differences in data samples. Data on life expectancy and gross national income per capita is taken from United Nations Development Programme’s website^7^. The measure of life expectancy is the ‘life expectancy index’ as available on the website. The complete data is available in Appendix A at the end of the article.

## Results

Table 1A presents the descriptive statistics of the data, and Table 1B presents the correlation matrix. The table replicates the results of Ilie and colleagues’ study. The coefficients of correlation between mean vitamin D level and Covid-19 cases and between mean vitamin D level and Covid-19 mortality are identical. The significance level is both cases is slightly higher than 0.05 (p value = 0.0501 and p value = 0.0535 respectively). However, as expected, life expectancy is strongly correlated with Covid-19 cases and mortality. Thus, it needs to be controlled for in modelling the impact of vitamin D levels on Covid-19 cases and mortality.

**Table 1A:**
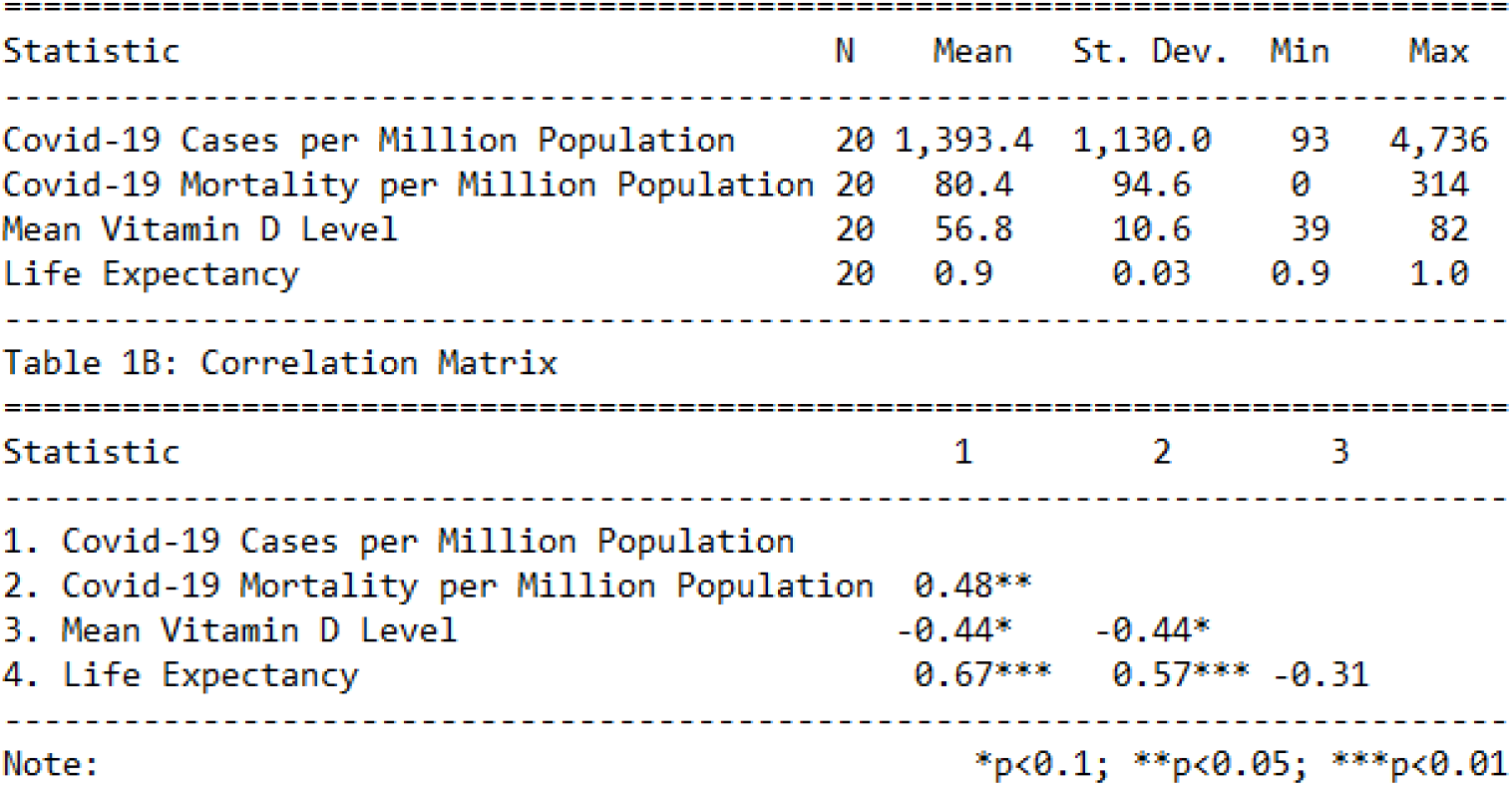
Descriptive Statistics

Tables 2A presents the results of linear regression model predicting Covid-19 cases per million population. Model 1 is a replication of the result in Ilie^4^ and colleagues’ study as it regressed mean vitamin D level against Covid-19 cases without any other variable. The coefficient is, as expected, negative and significant (β=-47.23, p<0.1). Model 2 regresses life expectancy against Covid-19 cases without any other variable. The coefficient in this case is positive and significant (β=23190.30, p<0.01). Model 3 includes both the variables of interest – mean vitamin D level and life expectancy – in the same model. The coefficient of mean vitamin D level is now not statistically significant (β=-27.65, ns). The coefficient of life expectancy is, however, still positive and significant (β=20394.39, p<0.1). This implies that vitamin D levels are not a predictor of number of Covid-19 cases at country level. Life expectancy is a better predictor as the coefficient of vitamin D loses significance in its presence in the model.

The positive coefficient of life expectancy might imply that countries with high life expectancy have better healthcare systems which in turn leads to better detection of cases. Another interpretation might be that countries with higher life expectancy have a more aged population. It is known that older people have higher chances of exhibiting severe symptoms of Covid-19. Severe symptoms are more likely to get tested, leading to better detection.

**Table 2A:** Results of Covid-19 cases per million population

|  | Dependent variable: |  |  |
| --- | --- | --- | --- |
|  | Covid-19 cases per million population |  |  |
|  | (1) | (2) | (3) |
| Mean Vitamin D Level | -47.231*<br>(22.496) |  | -27.650<br>(19.004) |
| Life Expectancy |  | 23,190.300***<br>(6,039.417) | 20,394.390***<br>(6,167.383) |
| Constant | 4,075.672***<br>(1,298.589) | -20,415.920***<br>(5,683.030) | -16,216.260**<br>(6,224.308) |
| Observations | 20 | 20 | 20 |
| R2 | 0.197 | 0.450 | 0.511 |
| Adjusted R2 | 0.152 | 0.420 | 0.454 |
| Residual Std. Error | 1,040.515 (df = 18) | 860.759 (df = 18) | 835.238 (df = 17) |
| F Statistic | 4.408* (df = 1; 18) | 14.744*** (df = 1; 18) | 8.888*** (df = 2; 17) |
| Note: *p<0.1; **p<0.05; ***p<0.01 |  |  |  |

Table 2B presents the results of linear regression model predicting Covid-19 mortality per million population. Model 1 is a replication of the result in Ilie and colleagues’ study as it regressed mean vitamin D level against Covid-19 mortality without any other variable. The coefficient is, as expected, negative and significant (β= -3.90, p<0.1). Model 2 regresses life expectancy against Covid-19 mortality without any other variable. The coefficient in this case is positive and significant (β=1650.69, p<0.01). Model 3 includes both the variables of interest – mean vitamin D level and life expectancy – in the same model. The coefficient of mean vitamin D level is now not statistically significant (β= -2.57, ns). The coefficient of life expectancy is, however, still positive and significant (β=20394.39, p<0.1). This implies that vitamin D levels are not a predictor of number of Covid-19 mortality at country level. Life expectancy is a better predictor as the coefficient of vitamin D loses significance in its presence in the model.

The positive coefficient of life expectancy might imply that countries with high life expectancy have better healthcare systems which in turn leads to better attribution of cause of death. Another interpretation might be that countries with higher life expectancy have a more aged population. It is known that older people have higher chances of developing fatal complications of Covid-19.

**Table 2B:** Results of Covid-19 mortality per million population

| Dependent variable: |  |  |  |
| --- | --- | --- | --- |
|  | Covid-19 mortality per million population |  |  |
|  | (1) | (2) | (3) |
| Mean Vitamin D Level | -3.904*<br>(1.890) |  | -2.568<br>(1.762) |
| Life Expectancy |  | 1,650.692***<br>(560.185) | 1,390.994**<br>(571.954) |
| Constant | 302.117**<br>(109.072) | -1,471.973**<br>(527.128) | -1,081.889*<br>(577.233) |
| Observations | 20 | 20 | 20 |
| R2 | 0.192 | 0.325 | 0.400 |
| Adjusted R2 | 0.147 | 0.288 | 0.330 |
| Residual Std. Error | 87.396 (df = 18) | 79.840 (df = 18) | 77.459 (df = 17) |
| F Statistic | 4.268* (df = 1; 18) | 8.683*** (df = 1; 18) | 5.674** (df = 2; 17) |
| Note: *p<0.1; **p<0.05; ***p<0.01 |  |  |  |

## Discussion

The results indicate that the correlation identified in Ilie and colleagues was most likely spurious in nature. The coefficient of vitamin D level lost its statistical significance in present of life expectancy. The life expectancy of a country was statistically significant predictor of both Covid-19 cases and Covid-19 deaths. The coefficient of life expectancy was positive in the models tested. This may be due to better healthcare systems which are better at detecting cases and attributing deaths to the disease. This might also mean that countries with higher life expectancy have higher proportion of older people. Although the results clearly indicate that vitamin D did not play a role in Covid-19 cases or deaths, a more definitive answer requires clinical trials. For now, the results indicate that vitamin D should not be seen as a cure of Covid-19.

This study can help avoid some of the overreaction that was seen in the case of HCQ despite weak medical evidence.

## Data Availability

Data of the study are publicly available.

## Appendix A

| *Countries* | *VitaminD* | *LifeExpectancy* | *CovidCases* | *CovidDeaths* |
| --- | --- | --- | --- | --- |
| *Norway* | 65 | 0.958 | 1123 | 19 |
| *Switzerland* | 46 | 0.979 | 2686 | 103 |
| *Ireland* | 56.4 | 0.955 | 1230 | 48 |
| *Germany* | 50.1 | 0.941 | 1309 | 25 |
| *Island* | 57 | 0.967 | 4736 | 18 |
| *Sweden* | 73.5 | 0.964 | 834 | 68 |
| *Netherlands* | 59.5 | 0.956 | 1199 | 131 |
| *Denmark* | 65 | 0.935 | 933 | 38 |
| *Finland* | 67.7 | 0.95 | 449 | 7 |
| *UK* | 47.4 | 0.942 | 895 | 105 |
| *Belgium* | 49.3 | 0.946 | 2019 | 193 |
| *Spain* | 42.5 | 0.976 | 3137 | 314 |
| *France* | 60 | 0.962 | 1671 | 167 |
| *Czech Republic* | 62.5 | 0.911 | 488 | 9 |
| *Italy* | 50 | 0.975 | 2306 | 292 |
| *Estonia* | 51 | 0.901 | 893 | 18 |
| *Slovakia* | 81.5 | 0.883 | 125 | 0.4 |
| *Portugal* | 39 | 0.952 | 1289 | 37 |
| *Hungary* | 60.6 | 0.872 | 93 | 6 |
| *Turkey* | 51.8 | 0.884 | 453 | 10 |

Note: CovidCases are number of Covid-19 cases per million population reported in each country. CovidMortality is number of deaths attributed to Covid-19 per million population in each country. Vitamin D is Vitamin D(25)HD mean measured in nmol/L. Life Expectancy Index data are taken from United Nations' Development Program website.

